# The experience and perception of first language usage in healthcare: the Welsh perspective

**DOI:** 10.1101/2025.02.20.25322596

**Authors:** Maisie E Edwards, Owen Bodger, Menna Brown, Llinos Roberts, Luke D Roberts, Jeffrey S Davies, Alwena H Morgan

## Abstract

Several studies have shown that using language that is most suitable for the patient is essential for effective communication in healthcare. Using a patients second language during health consultations can negatively impact a patient, causing delays in treatment and misdiagnosis. In Wales both Welsh and English have equal status in public sector organisations, however, independent primary care providers such as General Practices (GPs), do not need to comply with all the Welsh Language Standards. Thus, there is inconsistency in the availability of bilingual healthcare provision. This mixed methods study used a focus group and a survey of the Welsh speaking general population (361 participants) to gauge awareness of the Welsh Language Standards and collect experiences of bilingual healthcare, concentrating on GPs. The data underwent both qualitative thematic and quantitative analysis and revealed low awareness of the Welsh Language Standards (27%). Overall, respondents felt that their need to use Welsh is not taken seriously, with 71% having never been offered a Welsh consultation. 57% that have an English medium doctor reported that they would feel more comfortable having their consultations in Welsh. 32% of respondents from higher percentage Welsh speaking areas have felt restricted by their inability to communicate in their first language during GP appointments. There was overwhelming support for recording a patient’s preferred language on health records. The results suggest that the Welsh speaking public both want and need Welsh provision in primary care. However, there is a need to review primary care policies to facilitate a more effective roll out of an ‘active offer’ of first language healthcare in Wales.

## Introduction

Effective communication is an integral part of quality healthcare, however, the impact of the lack of bilingualism during healthcare provision has not been fully established. Linguistic barriers can compromise a patient’s healthcare opportunities (1). More specifically, ineffective communication in healthcare through the use of a patient’s second language can lead to misinterpretation of information and advice, less treatment compliance, poor psychological support (2) and a lack of informed consent (3). Furthermore, language discordance may lead to delayed treatment, misdiagnosis, longer hospital stays, medication errors, poor control of chronic conditions, increased risk of adverse events, or death (4–12)Notably, these effects have also been reported in studies concentrating on the effect of language discordance with vulnerable groups such as children (13–18) and older people, particularly in the context of dementia (19). Cultural differences between a healthcare physician and a patient can also lead to a lack of trust and contribute to adverse effects for the patient, even if they live in the same country (7,20–23).These reports largely come from a range of international studies investigating bilingualism/minority languages in healthcare settings, but very few have been carried out in the context of Welsh-medium Healthcare.

According to a census carried out in 2021, 29.1% of the population in Wales speak Welsh. Whilst all of the Welsh speaking population are considered bilingual (able to speak both Welsh and English), a survey by the Welsh consumer council reported that bilingual service users show a particular language preference during situations of stress and when they feel vulnerable (24). Furthermore, an individuals’ health literacy is lower in their second language (25). However, due to the status of Welsh as a minority language, there are fundamental deficiencies in Welsh-medium healthcare services, which could be disadvantageous to bilingual service users (24,26). Language choice is even more important for vulnerable bilingual patients, for example, children, who have not yet learnt English, people with dementia-related conditions who have lost their ability to speak English (their second language) (19) and people with learning disabilities or mental health disorders (27), who may find it easier to communicate and express themselves in a particular language. Restricted access of healthcare services to these cohorts in their preferred language has been shown to compromise the delivery of care (28). People in these groups who have lower levels of health literacy could be even further impacted by a lack of bilingual healthcare.

Since 2018, all Health Boards in Wales must abide and report on their compliance to the Welsh Language Standards (WLS), set by the Welsh Language Commissioner. However, the WLS do not apply to independent healthcare providers, such as general practice (GP) surgeries, as they are considered to be private businesses (29). Thus, apart from a small number of surgeries that are directly managed by health boards, the majority of GP surgeries in Wales do not have to provide any bilingual services or comply with the WLS. Furthermore, in these primary healthcare settings, patients do not have the right to demand the use of the Welsh language. Herein, we investigated the perceptions of the Welsh speaking public in relation to the use of Welsh in primary healthcare services and assess the demand/need for more bilingual rights and services in primary healthcare settings.

## Methods

Ethical approval was granted by Swansea University Medical School Research Ethics committee, (reference number 2021-0089, 18/11/2021). This study was carried out using a mixed methods approach. Stage 1 involved a review of the literature to inform the initial design of the questionnaire. This was supplemented by one focus groups to explore the experiences of people in Welsh in healthcare. Participants were identified through purposeful sampling. Known contacts were invited by email to take part in focus group discussions, to explore their views and experiences on the use of the Welsh language when receiving healthcare. These were facilitated via Zoom, between 01 December 2021 and 31 January 2022. All participants were first required to read a participant information sheet and provided written informed consent prior to taking part. Focus groups were audio and video recorded and transcribed by the researcher for analysis. The focus group data underwent qualitative thematic analysis (30) using nVivo. This followed a staged approach staged process was undertaken 1. familiarisation 2. initial coding was undertaken in the following way: the researchers (ME, AM) independently read, re-read and coded focus group one and two. Codes identified independently were shared and discussed and compiled into a coding structure document. Agreed codes were then applied to the remaining transcripts. New codes were shared and discussed as they emerged 3. The final codes were independently organised into themes. Themes were discussed until agreement was reached. Subthemes were created during this iterative process. Finally, 4. theme names were given, and extracts identified for each theme. A final review of each theme was undertaken to ensure no data were missed and coded data aligned with themes. Final themes were discussed with the research team and informed the survey design.

Stage 2 involved the roll out of the anonymous questionnaire which was prepared and hosted online using Qualtrics XM. The questionnaire was promoted to Welsh speakers within Swansea University and to the Welsh speaking public via known personal contacts as well as on social media (Twitter and Instagram). Inclusion criteria included being Welsh speaking, >18 years old, able to give informed consent and having access to a device with WIFI connection. Participants were first required to read the participant information sheet before consenting to commencing the survey. The questionnaire itself contained 28 questions, including demographics and language ability/preference of the respondents, followed by a series of questions relating to one of three key themes determined following discussions with the research team; 1) Awareness and opinion of the Welsh language rights provision in Healthcare; 2) Personal experiences and opinions on the use of Welsh in Primary care and 3) Impact of Welsh on the quality of care (Table 1). The questions were a mix of closed and open and free text questions where respondents were given the opportunity to expand on their thoughts/experiences. When firstly analysing the data, the region from where the participants originated seem to impact the results. Thus, in addition to analysing the data as one cohort we also carried out regional analysis. For regional analysis, the population was split into two, areas where more than or less than 35% of the local population speak Welsh (‘Higher’ Welsh speaking areas included: Gwynedd, Anglesey, Carmarthenshire, Ceredigion, Denbighshire and Conwy). This particular percentage was chosen as there seemed to be a natural divide in the percentage of Welsh speakers in each Welsh county at this level (Office for National Statistics, 2021). The data underwent quantitative analysis using SPSS (version 28) using Chi Square analysis unless stated otherwise with significance being indicated by a 5% change. The free text entries underwent qualitative thematic analysis using a pragmatic content analysis (31,32), informed by the results from Stage 1 and Stage 2. This was conducted by the research team (ME,AM). For some, question frequency analysis was then undertaken to identify the most commonly occurring themes.

**Table 1.**
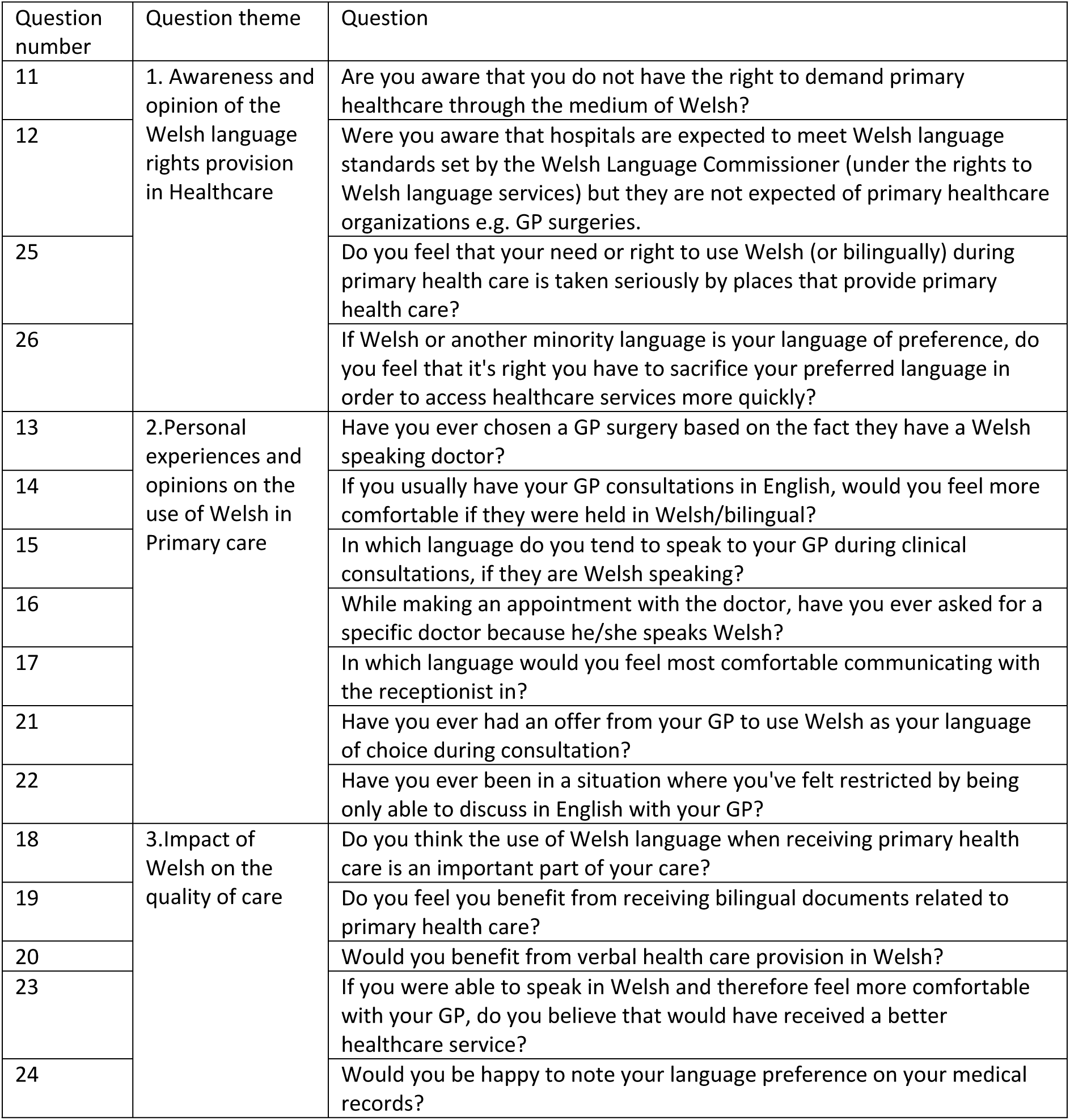
Questions with associated theme.

## Results

### General themes emerging from the focus group & dual interview

The inclusion criteria for the focus groups included being >18 years and Welsh speaking. The Participants included one >60 years old (male), two between 25-59 years old (two female) and one <25 years old (male). Due to sudden unavailability of two participants, a second focus was carried out as an extended dual interview (one participant >60 years old (male) and the other being 25-59 (female). The focus group and interview gave us the opportunity to explore people’s experiences to further inform the questionnaire. The transcript analysis revealed seven key themes, namely, ‘recognising the challenges that are facing the NHS’, ‘identification of vulnerable groups’, ‘the patient’s priorities’, ‘respect’, ‘the responsibility of the health carer to ensure that the patient understands’, ‘the use of people as unofficial translators’ and ‘the effect of using a patients first language’ (Table 2).

**Table 2.** Themes and sub-themes emerging from Focus Group with their associated quotations.

| Emerging Theme | Sub-theme | Example of associated quotation |
| --- | --- | --- |
| Vulnerable groups of people | 1.The Elderly | "Pobl uwchben wythdeg mlwydd oed, mae'r rhyddhad ma nhw'n cal pan ti'n mynd atyn nhw a phan ti'n siarad Cymrag da nhw, ti'n gweld e, ma fe jyst yn 'diolch byth', ma nhw'n gallu ymlacio. Mae'n rili bwerus. Fi'n cal hwna bron bob dydd. Fi'n cal 'diolch byth', 'diolch byth' bo chi'n siarad Cymraeg. A fi di cal pobl yn crio, yn dala'n ddwylo i, yn gweud 'oh doctor'. Mae'n neud shwd gymaint o wahaniaeth iddyn nhw, wir, mae'n rili rili bwysig i rai bobol."<br>*"People over eighty years old, the relief they feel when you go up to them and when you speak in Welsh, you see it, it's just 'thank goodness', they can relax. It's really powerful. I get that almost every day. I get 'thank goodness!, 'thank goodness that you speak Welsh'. And I've had people crying, holding my hands, saying 'oh doctor'. It makes such a difference to them, honestly, it's really important to some people."* (Female, 40-49) |
|  | 2. Children | "plant a henoed ydy'r ddau categori pwysicaf dwi'n meddwl da ni angen y gwasanaeth i fod yn Gymraeg achos, ti isio nhw ddallt yn does? deall be sy'n mynd ymlaen. Ma nhw'n teimlo'n 'vulnerable', ma nhw'n teimlo ddim yn cant y cant. So beth ma nhw angen ydy mynd at rhywun a nhw'n deud oce, paid a phoeni dyma beth sydd yn mynd ymlaen, dyna beth da ni angen, dyna beth ydy dynolaeth, dyna pam da ni'n mynd at ddoctoriaid de, iddynt nhw ddaethon ni, oce newn ni sortio fo."<br>*"children and the elderly are the two most important categories that I think we need the service to be in Welsh, because, you want them to understand don't you? to understand what's going on. They feel vulnerable, they don't feel 100 percent. So what they need is to go to someone and they say ok, don't worry this is what's going on, that's what we need, that's what humanity is, that's why we go to doctors, ok we'll sort it out."* (Female, 20-29) |
|  | 3.People with mental health issues | "fi di cal cleifion ifanc, iau na fi, pobl yn ei ugeinie pan oni yn y Feddygfa yn dod ata i, a fi'n cofio un yn benodol, odd hi'n diodde o iselder ysbryd a bob tro odd hi'n dod mewn i'r ystafell odd hi'n gorjys, pob tro odd hi'n dod mewn odd hi'n dweud 'fi'n dod ato ti Dr *** achos fi'n gallu siarad da ti, fi'n gallu siarad Cymrag. Fi ffael siarad da neb arall....mae'n effeithio pobl ifanc hefyd."<br>*I've had young patients, younger than me, people in their twenties come to me when I was at the Surgery, and I remember one in particular, she was suffering from depression and every time she come into the room she was gorgeous, every time she came she'd say 'I come to you Dr \*\*\*\* because I can talk to you, I can speak in Welsh . I can't speak with anyone else'.... it affects young people too."* (Female, 40-49) |
| Effect of using a Patients' first language | 1.Comfortability | "Ma fe'n anodd achos ma lot mwy gyfforddus siarad yn y Gymraeg na beth yw e yn y Saesneg, yn enwedig os ma fe wrth glywed newyddion chi ddim isie clywed. Lot yn neisach clywed e yn iaith dy hun. Dim ond unwaith fi wedi gorfod gweld meddyg yn [lleoliad]. Odd neb yn siarad Cymrag lawr fan hyn."<br>*"It's difficult because it's a lot more comfortable to speak in Welsh than what it is in English, especially if it's news you don't want to hear. Much nicer to hear it in your own language. I have only had to see a doctor in [location] once. No one speaks Welsh down here."* (Male, 20-29)<br>"Ond mynd yn ôl at yr iechyd meddwl, pan ma pobl yn 'vulnerable', yn fregus, pan ma rhywun yn fregus ti'm isio gorfodi nhw i gorfod, wel sygyron nhw ddim yr egni i feddwl am yr eirfa mewn iaith arall de. Ti isio nhw fod mor gyffyrddus ag sy'n bosib, so bod nhw'n rili gallu agor i fyny a deud dyma beth sydd yn mynd ymlaen, ac os wyt ti'n gorfod meddwl am iaith a hyn hyn a llall, ti falle ddim yn mynd i gal y llun cryfa" /<br>*"But going back to mental health, when people are 'vulnerable', frail, when someone is fragile you don't want to force them to have to, well they don't have the energy to think about the vocabulary in another language right. You want them to be as comfortable as possible, so that they can* |
|  |  | *open up and say this is what's going on, and if you have to think about language and this and the other, you might not get the best picture"* (Female, 20-29) |
|  | 2.Expression of the patient | *"Mae fel iaith wahanol, mae fel iaith ffurfiol da chi ond yn defnyddio pan da chi'n sal, ond wrth gwrs pan da chi'n sal da chi ddim eisiau poeni lot am bethau so mae'n rhaid paratoi ar gyfer y sefyllfa na rhywsut." / "It's like a different language, it's like a formal language you only use when you're ill, but of course when you're ill you don't want to worry too much about things like this, so you have to prepare for the situation no somehow."* (Male, 70-79) |
|  | 3.Developing a rapport with a patient | *"o bwynt gwneud rappour, o'r foment gynta, bydden i yn gweud ie, mae yn dylanwadu fi'n credu beth i chi'n meddwl o'r person o'ch blaen chi." / "from the point of making rappour, from the first moment, I would say yes, i think it influences what you think of the person in front of you."* (Female, 40-49) |
| A patient's priorities | 1. Quality of healthcare | *"ydy'n ni'n gorfod aberthu un am llall bron, de? dyle ni ddim gorfod, dyna eto ydy'r ideal senario, bod ni'm yn gorfod. A bod gynnon ni ffydd yn yr Cymry, bod gynnon ni ddoctoriaid, y gore, ar gael i ni. Ond dwi'n meddwl pe bydda yr hawliau yma yn dod i fyny, fysai o bosib, dwi'n trio meddwl o bob persbectif o fama, fysa o bosib rhai teuluoedd, neu siaradwyr Cymraeg bron ddim yn hawlio achos fysa nhw isio'r doctor gora" / "do we have to sacrifice one for another almost, right? We shouldn't have to, that is again the ideal scenario, that we don't have to. And that we have faith in the Welsh doctors, that we have the doctors, the best, available to us. But I think that if these rights come up, it would be possible, I'm trying to think from every perspective here, it could be that some families, or Welsh speakers don't claim the right because they want the best doctor"* (Female, 20-29) |
|  | 2.The importance of health | *"beth dwi'n trio pwysleisio ydy yn enwedig pan mae o'n dŵad i iechyd, iechyd sydd yn bwysig"/ "What I'm trying to emphasize is especially when it comes to health, health is important"* (Female, 20-29) |
| The responsibility of the health carer to ensure that the patient understands |  | *"Fedri di sefyll fana a deud y sbiel efo'r geiriau mawr ma, grêt, ond efo doctoriaid mae o'n gyfrifoldeb arna nhw i sicrhau bod y patient yn deall beth sy'n mynd ymlaen er mwyn tynnu'r lefelau straen na, os ma nhw mewn cyfnod ofnadwy weithiau ma nhw'n rili... mae o'n bywyd neu marwolaeth i rai sefyllfaoedd, ti eisiau neud y patient sydd gynaf, ac er mwyn neud yn siŵr bod y patient sydd gyntaf, mae'r rhaid neud yn siŵr bod nhw'n deall bob elfen o'r sefyllfa a neud iddyn nhw teimlo'n hollol gyfforddus. I lot o bobl, yn amlwg yng Nghymru, Cymraeg ydy'r ffordd o neud hynny achos mamiaith ydy o, y gwreiddyn tyfna sydd gynno ni, so mae'n rhaid i ti gysylltu efo hyna er mwyn gwneud i bobl teimlo'n gartrefol" / "You can stand there and say the spiele with these big words, great, but with doctors it is their responsibility to ensure that the patient understands what is going on in order to remove the stress levels, if they are in a terrible phase sometimes they are... it is life or death for some situations, you want to put the patient first, and in order to make sure that the patient comes first, you have to be sure they understand every element of the situation and make them feel completely comfortable. For a lot of people, obviously in Wales, Welsh is the way to do that because it's their mother tongue, we have deep root there, so you have to connect with that in order to make people feel at home"* (Female, 20-29) |
| Use of unofficial translators |  | *"mae o wedi sylwi, mae o yn gorfod ailadrodd a chyfieithu beth mae'r doctoriaid ma'n ddeud i'r pobl sydd yn aros yma, y patient, mae o'n gorfod, ti'n gorfod mynd at lefel y patients yndwyt, os dydy nhw ddim yn deall yna wel beth ydy'r point." / "he has noticed, he has to repeat and translate what the doctors are saying to the people who are staying here, the patient, he has to, you have to go to the level of the patients don't you if they don't understand, well what's the point."* (Female, 20-29) |
| Respect |  | *"fi'n credu taw parch yw e. Nid falle, odi e'n neud i chi deimlo'n fwy cyfforddus yn syth, dim rili i fod yn hollol honest. Bydden i yn digon hapus, digon cyfforddus sa fe'n gweud 'hello, how are you?'. Os yw e yn gweud "Bore da", fi syth yn meddwl "Oh good egg", oleia' ma fe'n gwneud ymdrech. Ma fe'n parchu fi." /* |
|  |  | *"I think it's respect. Not that it makes you feel more comfortable straight away, not really to be completely honest. I'd be happy enough, comfortable enough if he said 'hello, how are you?'. If he says "Bore da (Good morning)", I immediately think "Oh good egg", then he making an effort. He respects me." (Female, 40-49)* |
| Recognition of the challenges facing the NHS | 1. Fears about the practicalities of bilingual provision | *"O'r safbwynt yn ochr gwladgarol, ma na fwy iddo fe na jyst byse fe'n neis i gael, achos ma na oblygiadau wahanol iddo fe hefyd. Ond da ni'n mynd i werthu'r syniad i neb, mae 'na gwestiwn ymarferol" /*<br>*"From the point of view of the patriotic side, there's more to it than it only being nice to have, because there are different implications for it too. But we're not going to sell that idea to anyone, there's a practical question" (Female, 20-29)* |
|  | 2. General lack of Healthcare workers, let alone ones that are bilingual | *"Mae'r ateb delfrydol yn amlwg, dim jyst am y Gymraeg, ond does dim digon o doctors a nyrsys" /*<br>*"The ideal solution is obvious, not just about the Welsh language, but there are not enough doctors and nurses" (Male, 70-79)* |
|  | 3. Problems with staff recruitment in addition to retaining staff | *"trwy uniaethu a Chymru, mi fysai yn datrys broblem o recriwtio a phwysicach na hynny, cadw yr staff yng Nghymru" /*<br>*"by identifying with Wales, it would solve the problem of recruitment and more important than that, keep the staff in Wales" (Male, 70-79)* |

### Questionnaire results

#### Data Summary

Of the 377 submitted forms 361 were sufficiently complete and included in the final data set. The sample were predominantly female (76%, 272/357) and mostly in the 30-49 age range (45%). Around a quarter were less than 30 or more than 50 (26% and 29% respectively). Respondents are drawn from across Wales with representatives of all 22 Welsh counties. The proportion from areas with higher levels of Welsh speakers (determined as being over 35% of the local population) was 44%. These areas included the counties of Gwynedd, Anglesey, Ceredigion, Carmarthenshire, Conwy and Denbighshire. The overwhelming majority were currently in work (73%), with students making up a further 16%.

#### Welsh language status of the participants

While the sample was recruited from those who are Welsh speaking, only 53% declared that Welsh was their first language at home, with 18% using both languages. This left approximately a quarter of respondents reporting English as their primary language at home. This figure varies significantly with age (p=0.004), with those over 50 being much more likely to speak Welsh at home (63%) compared to those under 30 (38%).

Most of the sample describe themselves as being comfortable using the Welsh language verbally (77%), in writing (60%) and when reading (61%). While age does not appear to affect comfort in writing and reading, there is a significant association (p=0.008) between age and spoken language, with half (50%) of the over-50s show a preference for expressing themselves verbally in Welsh.

Respondents from Welsh speaking regions are much more likely (p<0.001) to have Welsh as their first language at home in comparison to respondents from regions with less than 35% Welsh speakers (91% vs 56%). We see a similarly strong, and expected, result in expressing themselves in Welsh, with 91% (verbal), 71% (writing) and 75% (reading) either preferring to use Welsh or being equally comfortable in both languages.

##### Results Theme 1 – The awareness of the general public on the lack of right to access to bilingual primary healthcare in Wales

In 2022 the Welsh language Commissioner launched the ‘Mae gen i hawl’ (‘*I have a right’*) campaign which promoted awareness of the right of the public to Welsh-medium services. However, the Welsh language standards only apply to public entities such as universities and health boards and not private establishments. To further complicate the situation, whilst approximately 9% of GP surgeries are run by the local health boards (Welsh government statistics, 2019), the majority are considered private establishments. Thus, there is likely to be confusion regarding who do and do not have to offer bilingual services. From the questionnaire, the public seem largely unaware of their rights in this respect. Only 27% were aware that hospitals are expected to meet Welsh Language Standards in the delivery of bilingual services. A similarly small percentage (28%) are aware that the rights to bilingual services do not extend to independently run primary healthcare establishments (e.g. GP surgeries). Older people are significantly more aware of these rights, with the over 50s being twice as likely to know their rights compared to the under 30s. Awareness of language rights in hospitals among the over 50s is 37%, compared to 19% for the under 30s. The equivalent figures for language rights in independent primary care establishments were 36% and 17% respectively. We did not observe any significant variation in these rates by region or gender.

As a whole, the sample felt strongly that their need to use their first language is not taken seriously. Only 18% felt that this was given sufficient priority in primary health care. Furthermore, over half (53%) felt that it was wrong that they were required to give up the use of their preferred language in order to access healthcare services more quickly. Those living in Welsh speaking regions were much less more likely to say that their language needs were being taken seriously in primary healthcare (p=0.008), relative to those in other regions (27% vs 14%).

##### Result theme 2. Personal experiences and opinions on the use of Welsh in Primary care

Having assessed the general knowledge of the public on their rights to bilingual provision, we next wanted to investigate their opinion and personal experiences of bilingual healthcare services. Nearly half of respondents reported that they choose to converse with their GP in Welsh (44%, 156/354 respondents). If we consider only those that have a Welsh speaking GP, this figure rises to 60% (156/264). A further 13% (34/264) choose to communicate bilingually. We see a significant difference according to age (p=0.001), with the 82% (70/85) of the over-50s choosing to speak partly, or wholly, in Welsh compared to 52% (32/62) of the under 30s. In Welsh speaking regions the average figure, across all ages, is even higher at 89% (119/133) and even in the non-Welsh speaking regions over half choose to use Welsh to some extent (54%, 71/131) if the GP is able to speak in Welsh (p<0.001). Where patients usually use English with their GP, the majority (57%) report that they would be more comfortable if those conversations were in Welsh or bilingual. Even among the under 30s and in non-Welsh speaking regions these figures were 49% and 42% respectively. While the consultation is the most critical part of engagement with primary care the overwhelming majority (84%) would feel more comfortable if they were able to use Welsh (including bilingually) with the reception staff.

Despite the strong demand for the use of the Welsh language, 71% (246/346) reported not being offered the option. Given the high numbers who do use Welsh this suggests that the option is not always offered even where the GP can speak Welsh. However, Welsh provision is offered far more often (p<0.001) in Welsh speaking regions, with rates of 32% vs 14% for non-Welsh speaking regions.

Unsurprisingly, this means that a substantial minority of respondents (24%, 83/345) report having felt restricted by their inability to communicate in their preferred language. This was significantly higher in Welsh speaking regions (32% vs 18%). Considering the free text responses for this question, the majority of these experiences were in the context of children (47%) and the elderly (37%), with additional experiences in context of mental health (10%), dementia (3%) and maternity (3%).

This issue is taken very seriously by many, with 18% (63/354) specifically requesting Welsh speaking GPs and 20% (72/358) going so far as to choose surgeries that employ Welsh speaking GPs. This figure is highest in Welsh speaking regions with 25% (40/159) and 29% (46/159) actively choosing GPs or GP surgeries based on this fact. Among those with a strong preference for the use of Welsh during a consultation well over a third (41%, 64/156) have chosen a surgery where this will be possible.

##### Results theme 3-Impact on quality of care

The questionnaire then explored the patients’ opinion on the impact of Welsh provision on their quality of care. The overwhelming majority of those sampled (67%, 233/346) felt that they would benefit from verbal health care provision in Welsh. Older patients felt even more strongly (80%) but even among the under 30s the majority agreed (55%) (p<0.001). The effect per region was just as pronounced, with 81% in Welsh speaking regions clearly indicating they believed they would benefit. Even in non-Welsh speaking regions over a half (56%) shared this belief.

About a third of respondents (31%, 105/337) believed that being able to speak Welsh with the GP would enhance their treatment, with 38% (129/337) disagreeing. There was a much stronger response to the use of bilingual documents with 63% (220/350) reporting that this would improve their healthcare. This figure was higher (69%) in Welsh speaking regions (p=0.016). Twenty-two comments were left in the free text boxes for this question which revealed several themes. In terms of comments relating to there not being an enhancement in their treatment if it was delivered in Welsh, there were 5 main themes: ‘no improvement in services’ (3 comments); ‘patients were just as comfortable using English’ (3, comments), ‘a lack of availability of Welsh speakers at their practice’ (2 comments); ‘a restriction in services’ (1 comment) and that ‘there might be a better service but not care’ (1 comment). Conversely, there were 13 themes associated with there being a beneficial effect of their treatment being delivered in Welsh. These included ‘groups with increased requirement for the provision’ (6 comments), ‘expression by the patient’ (4 comments); ‘communication’ (5 comments); ‘comfort’ (2 comments); ‘improving the patient-clinician bond’ (2 comments); then one comment each in relation to ‘understanding’, ‘support’, ‘relief’, ‘being judged on their English’, ‘the explanations of the patient being disregarded by English speaking clinicians’, ‘a reduction in the linguistic barrier’, ‘difficulties when using translators’ and ‘improved quality of care’. There was also one comment that was associated with being unsure of the impact of receiving their treatment in Welsh.

It is clear from the responses as a whole that the Welsh language provision in Primary Care is an important issue. When asked directly whether use of Welsh language is an important part of their care when receiving primary healthcare, three quarters agreed (73%), with 44% stating that it was very important. In Welsh speaking regions this figure was higher with 89% respondents rating it as important, but even in non-Welsh speaking regions it was a majority position (61%).

In order to facilitate an ‘active offer’ of Welsh language provision in primary care (rather than being requested) the patients would need to make their preference clear on their medical records. We found almost complete support for this, with 86% of respondents being happy for a record of their language preference to be made and a further 14% being unconcerned by this. This left only one single individual in the survey that would not be happy with this.

#### Thematic analysis results of the questionnaire free text data

Many of the questionnaire participants left in-depth comments, suggesting that many felt passionate about this topic. Overall, there were 7 main emerging themes which included ‘Vulnerable groups of people’, ‘language choice’, ‘effect of using a patients first language’, ‘a patients’ priorities’, ‘the presumption that everyone can and is happy to communicate in English’, ‘current services’ and ‘recognition of the challenges facing the NHS’ (Table 3). 4 of these themes overlapped with themes from the focus groups.

**Table 3.**
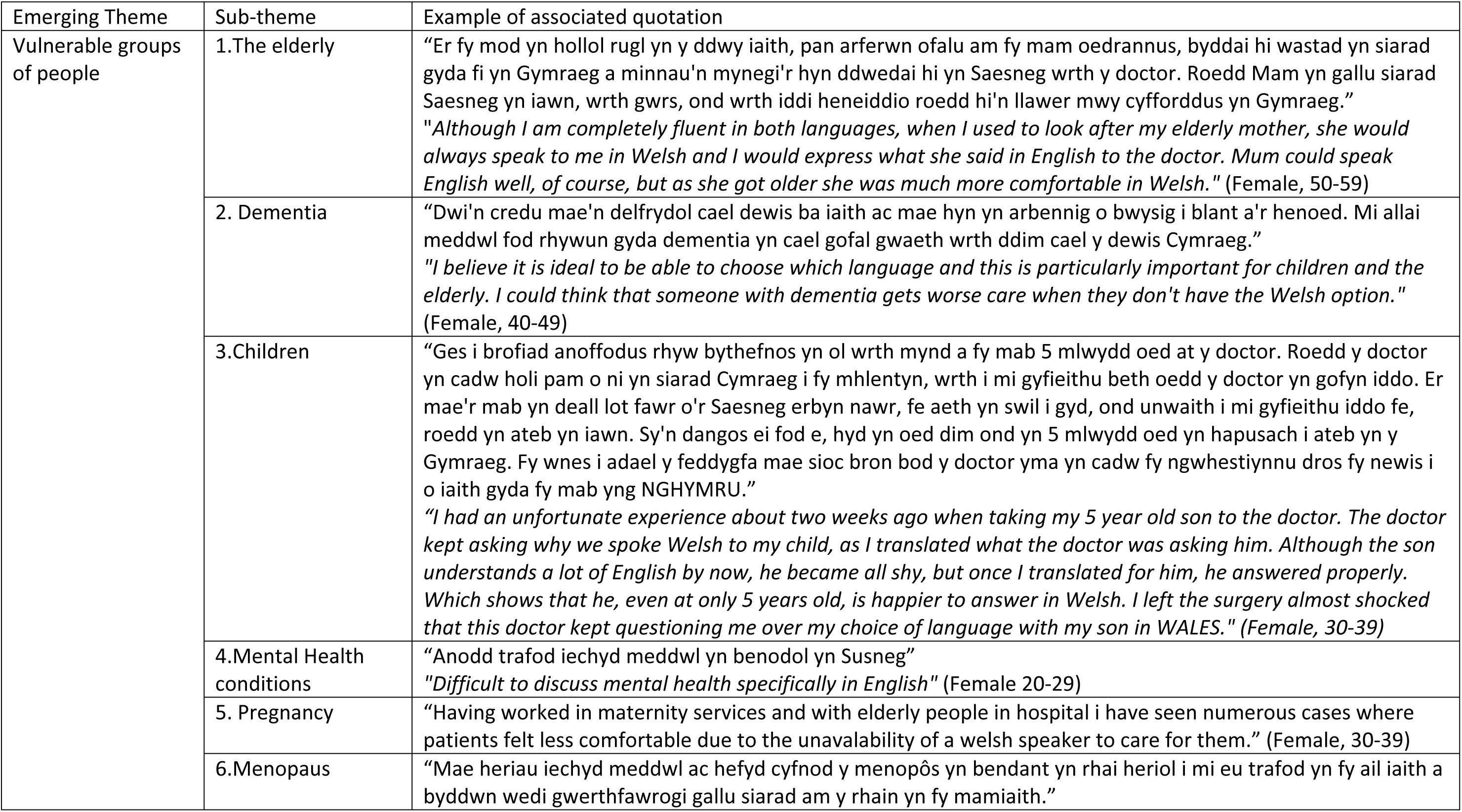
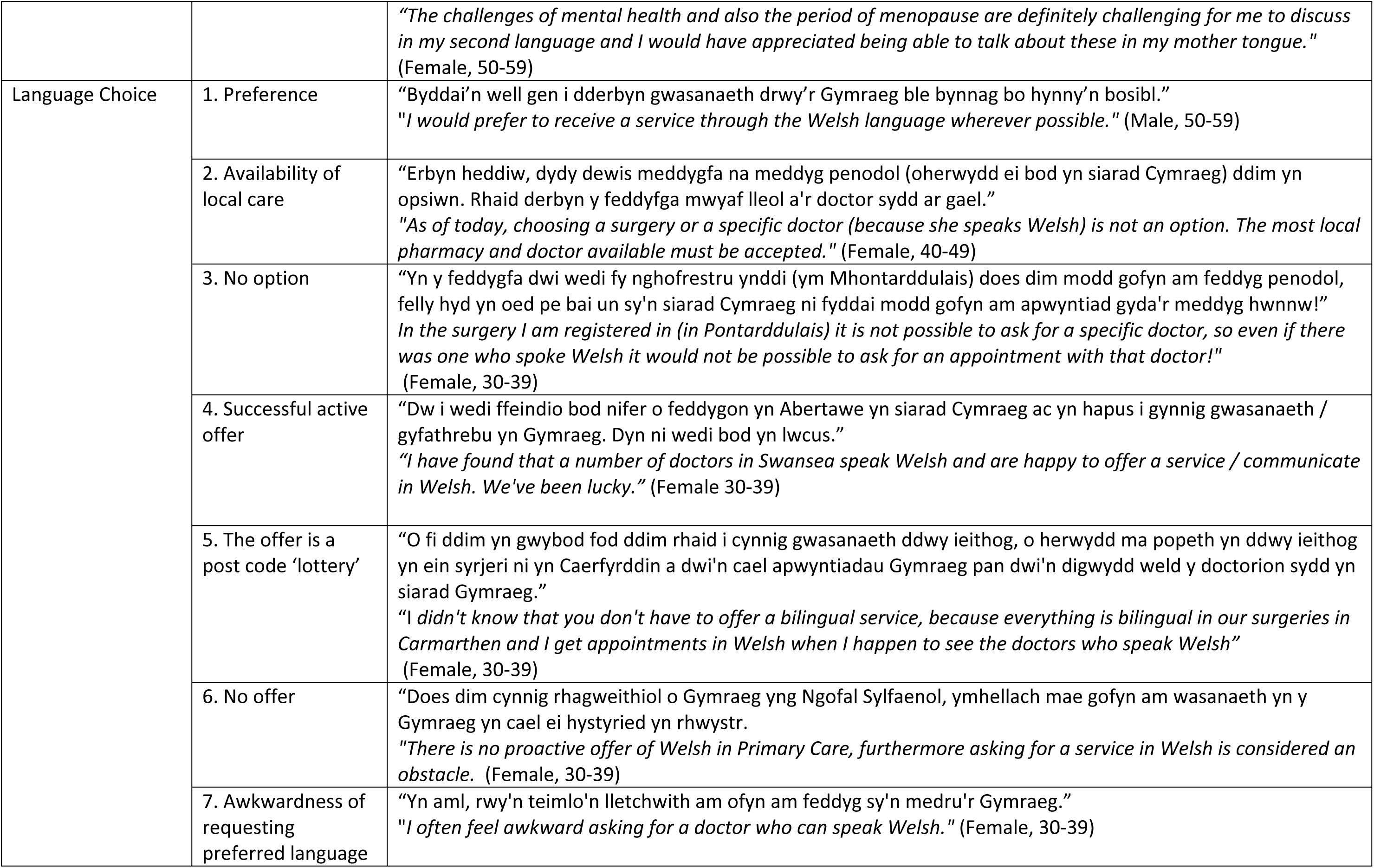
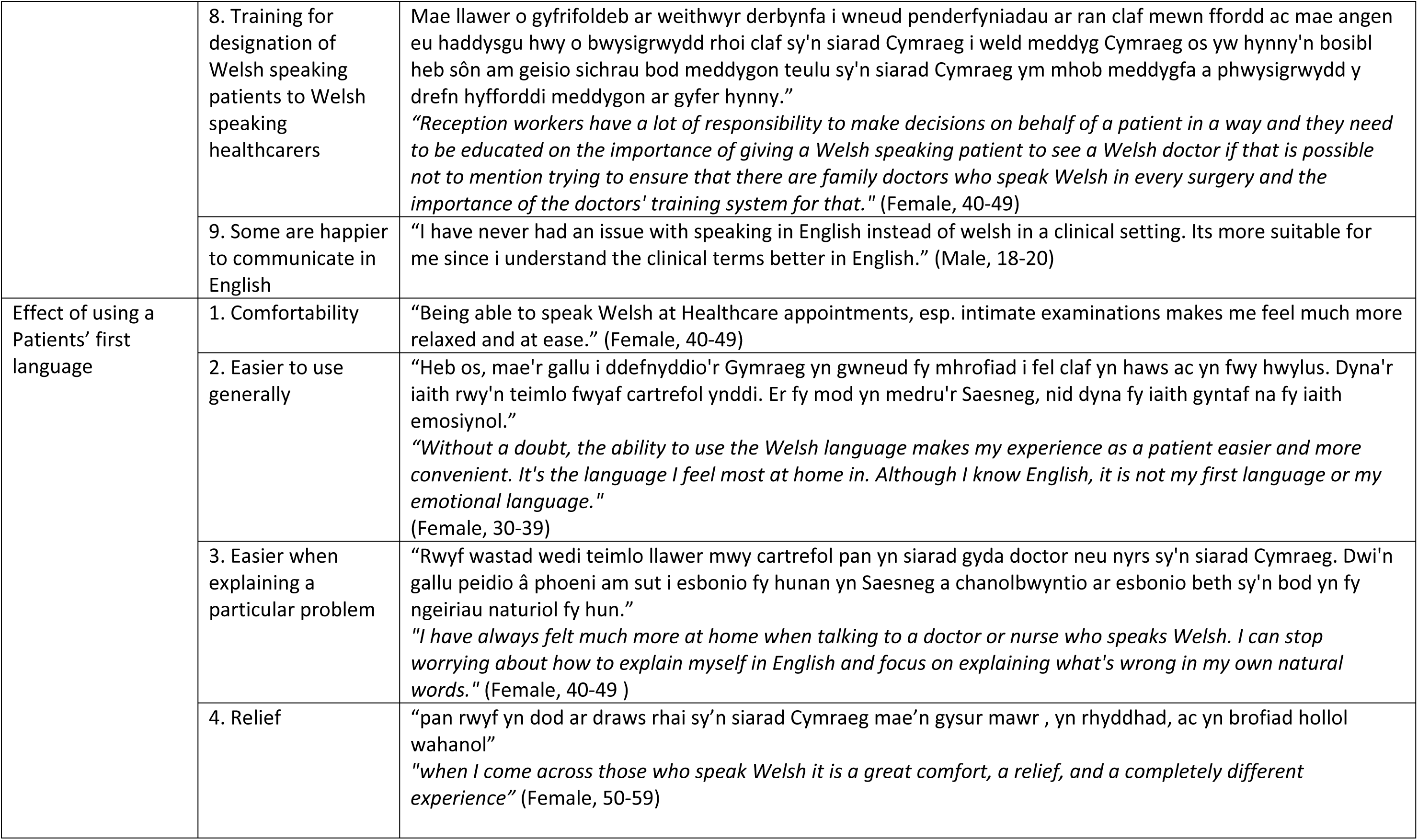
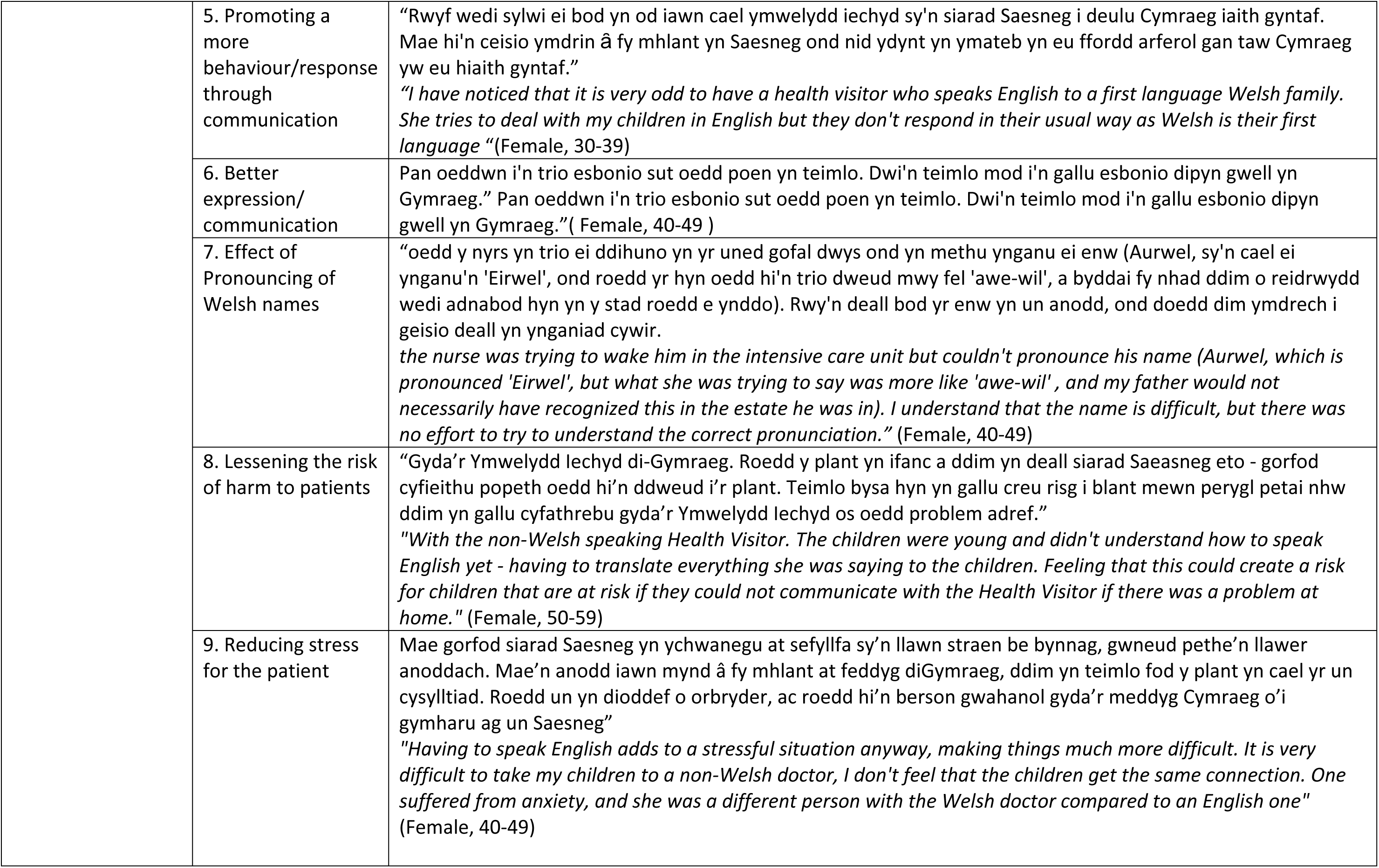
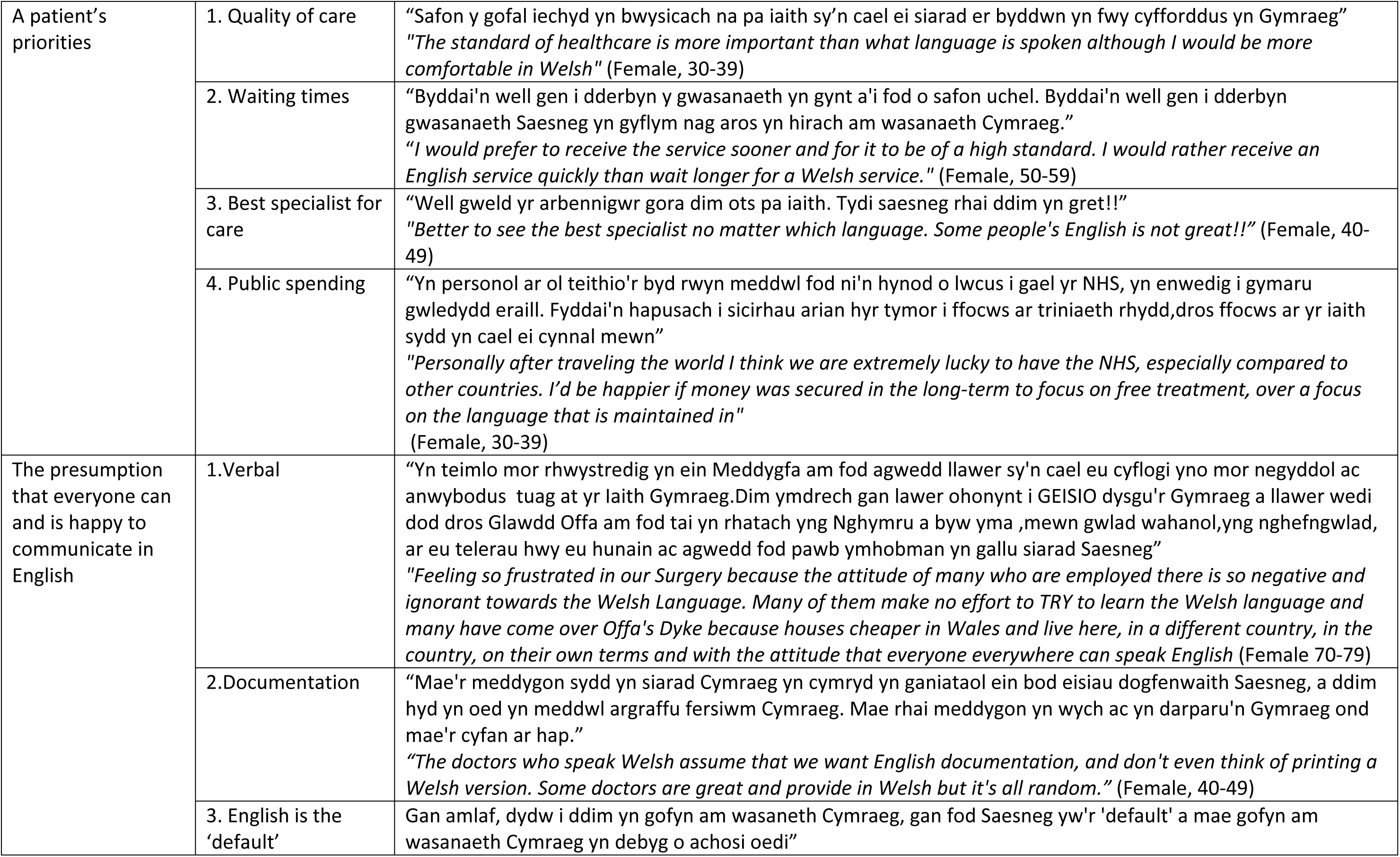
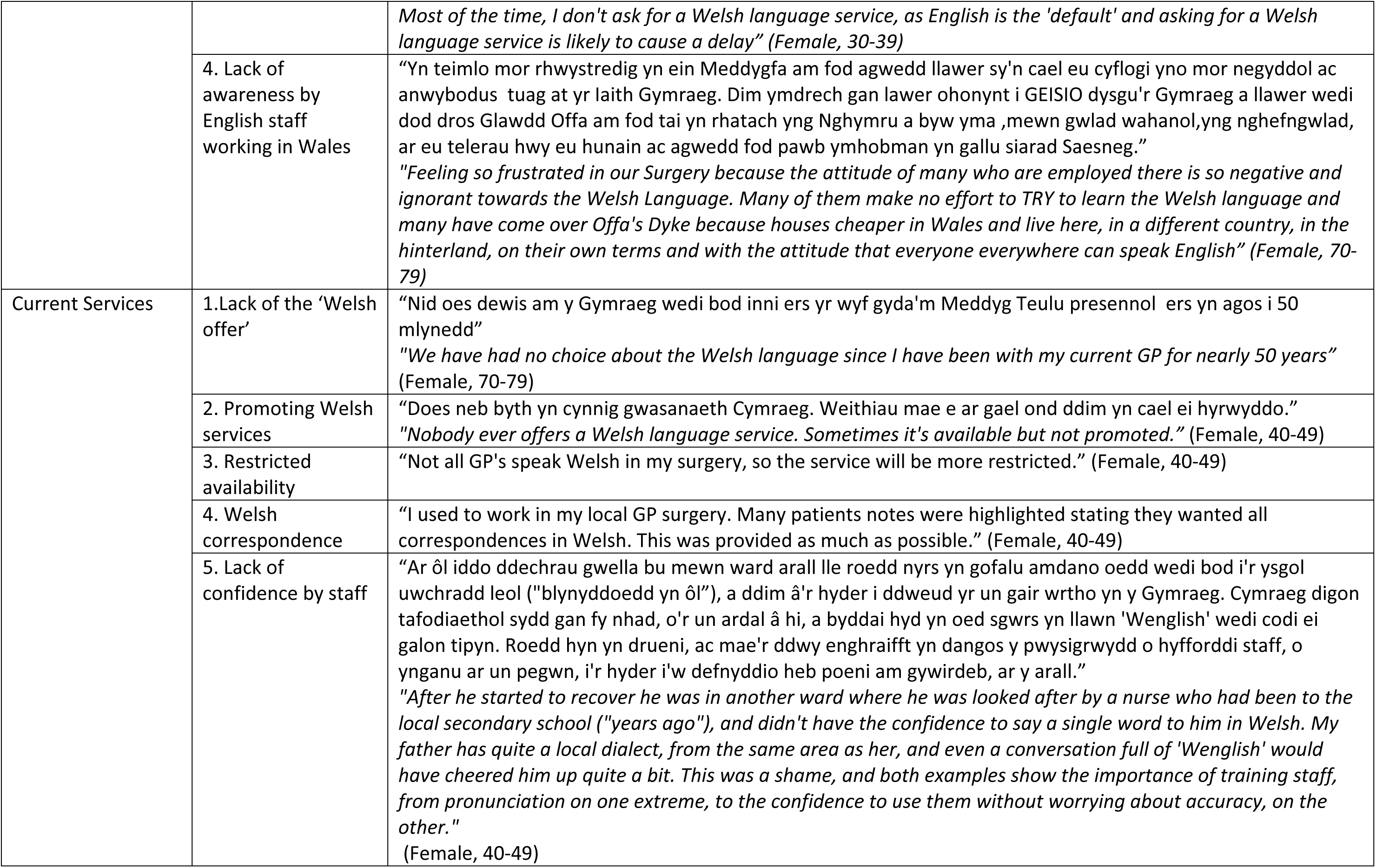
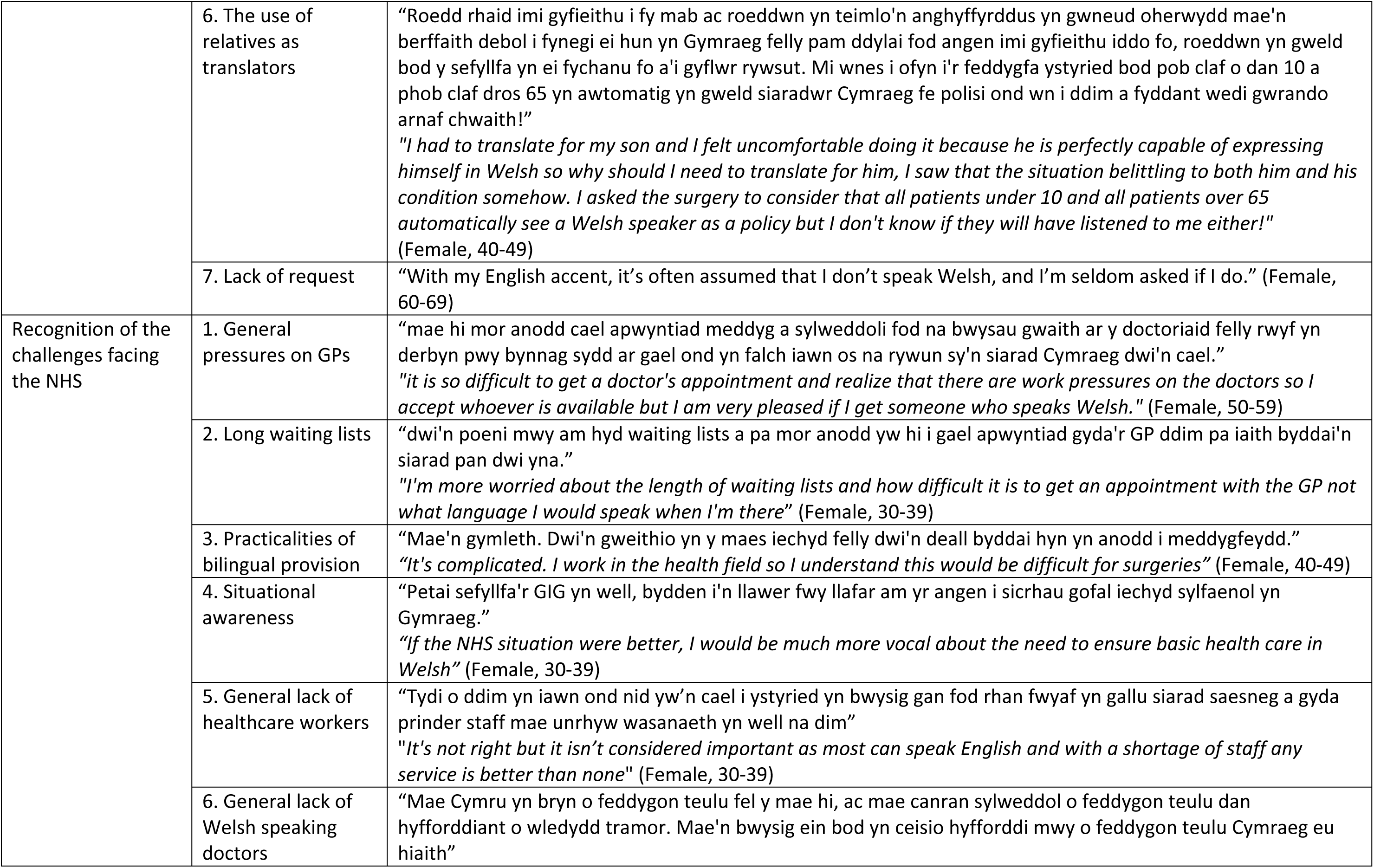
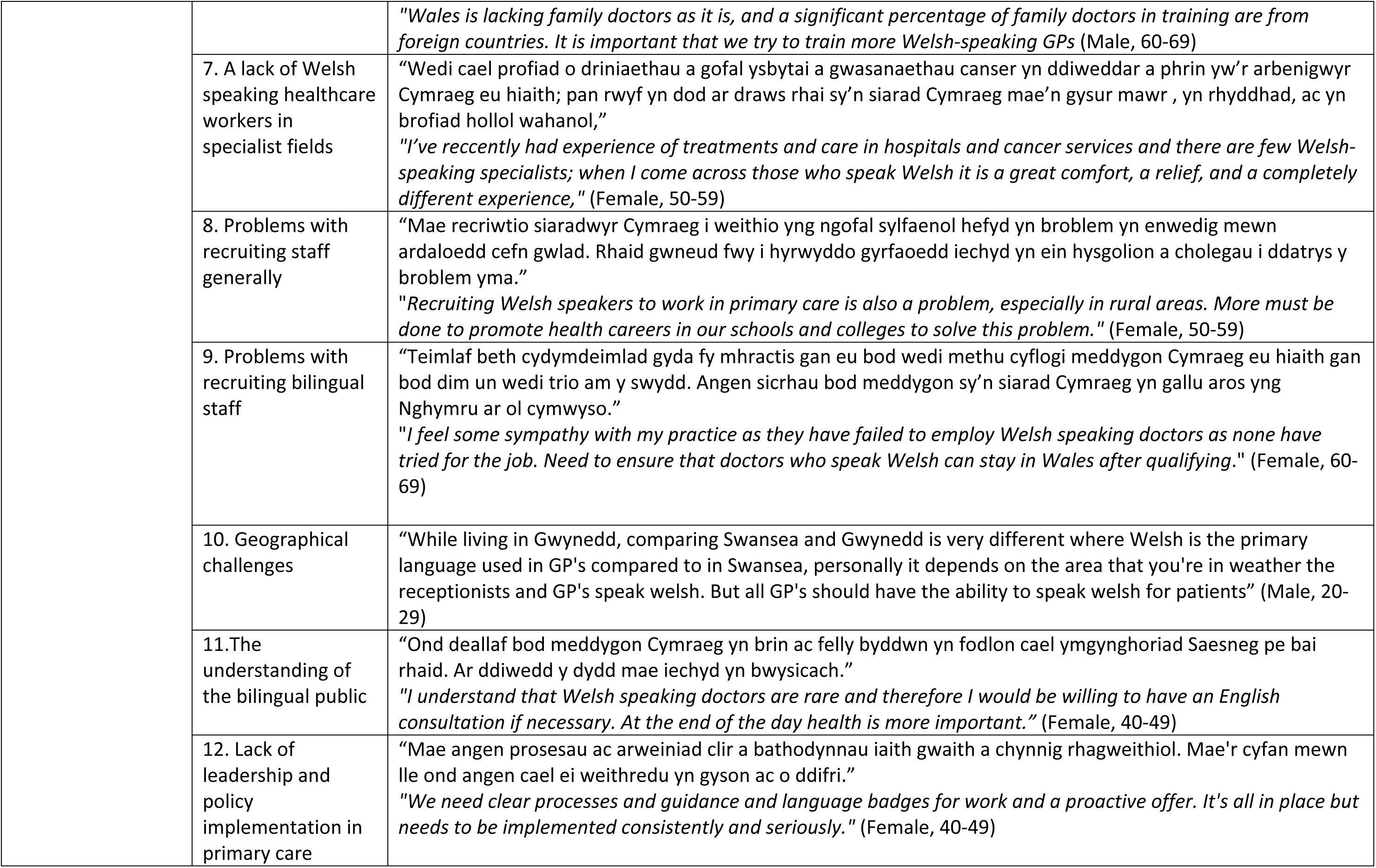
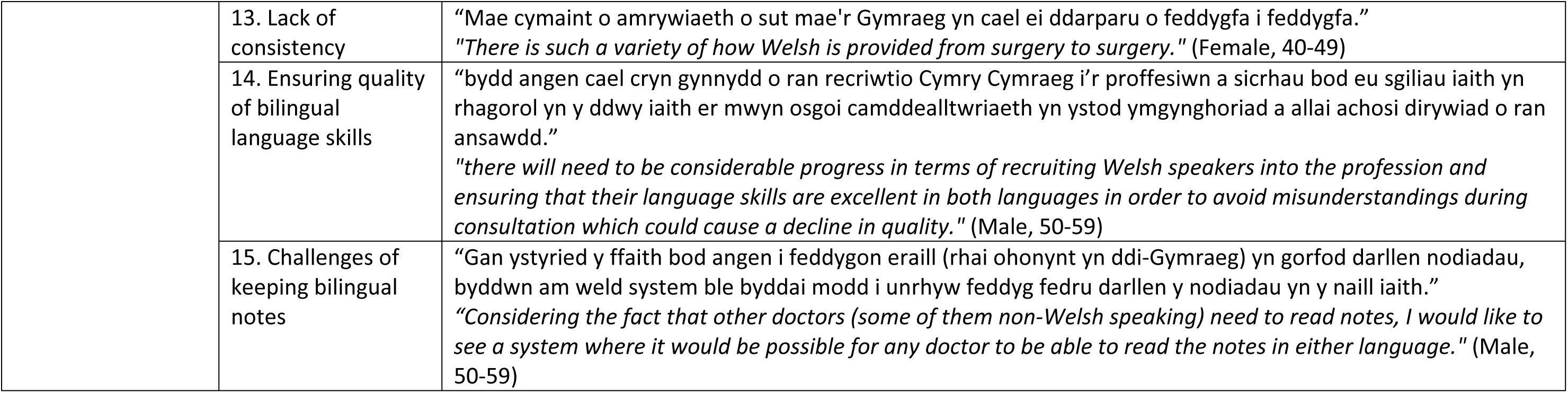
Themes and sub-themes emerging from free text boxes in the questionnaire with their associated quotations.

## Discussion

Whilst effective communication is important for quality healthcare, the impact of the lack of bilingual healthcare on first-language Welsh speakers has not been strongly established. This study used a focus group/extended dual interview to develop an online survey designed to explore the opinion of Welsh speakers on the use of Welsh in general practices (GPs). The questionnaire’s results revealed that 53% of participants spoke Welsh at home, with 18% using both Welsh and English. These figures were higher than those reported in the Welsh government’s ‘Welsh in the home’ survey (2022) (being 35% and 9% respectively) (33). However, this may not be surprising given that the questionnaire targeted Welsh speakers and there may be some responder bias as Welsh speakers might feel more inclined to participate in a survey of this nature. Whilst this is a potential limitation of the study it is still important and relevant to document the experiences of Welsh speakers.

When we assessed the awareness of Welsh speakers of their rights to bilingual care, only a third of respondents were aware of the WLS and their application to hospitals but not to most of GPs. Considering the WLS came into force in 2016, this highlights slow public information dissemination. This may suggest the need for a public campaign to clarify rights and expectations in Welsh healthcare settings.

When we investigated opinions on the use of Welsh in primary care there was consistently a more pronounced difference in the answers from people residing in higher percentage Welsh speaking regions. For example, those living in higher Welsh-speaking regions were more likely to perceive benefits from Welsh verbal healthcare provision. Furthermore, they were more likely to choose a GP with Welsh provision. This may be as expected due to a general assumption and expectation of available bilingual provision within these higher percentage Welsh areas. However, bilingual availability often resembles a post code lottery, as noted by one respondent, *“No one ever offers a Welsh language service. Sometimes it’s available but not promoted”.* This observation raises concerns that some services may exist but go underutilized due to a lack of active promotion or visibility. For many, the preference extends beyond speaking Welsh with a GP, 84% of respondents expressed feeling more comfortable using Welsh with the reception staff. The *Iaith Gwaith* (Working Welsh) logo, launched in 2005 by the Welsh Language Commissioner, is designed to indicate Welsh-speaking staff through an orange speech bubble/apostrophe symbol (’). However, the logo’s promotion and consistent use in GP surgeries appear limited, and its meaning may not be widely understood by the public. Raising awareness and encouraging the use of the *Iaith Gwaith* logo could empower patients to identify and request services in their preferred language without relying solely on an ‘active offer’.

Notably, only 18% of respondents felt their need to use Welsh was taken seriously in healthcare settings. One respondent reported “*The doctor kept asking why I spoke Welsh to my child, as I translated what the doctor was asking him…I left the surgery almost shocked that this doctor kept questioning me about my choice of language with my son in WALES” (Female, 30-39)*. Communication with patients is an important aspect of primary healthcare provision. The General Medical Council 2013) emphasizes that doctors have a duty to provide information to patients ‘in a way they can understand’. For Welsh-speaking patients, using their first language can be important in achieving effective communication, particularly in vulnerable situations, such as when dealing with children. Several respondents highlighted how using Welsh as their first language aids in explaining their problems, in addition to promoting comfort and relief (Tables 2 & 3). This is consistent with the literature which suggests that in stressful or uncertain situations, Welsh speakers feel more confident communicating in their first language (1,24). International evidence also supports the significance of first-language care. In the United States, patients’ fears are alleviated when doctors speak to patients through their first language (34). Furthermore, in Finland it was found that patients feel safer and have a greater sense of trust when they receive care in their first language whilst language discordance exacerbates symptoms (35). Indeed, language concordance between patients and healthcare providers can help build stronger relationships, encouraging patients to share more information. This can lead to more accurate diagnoses and improved outcomes (36). These findings underscore the importance of providing bilingual services in primary care to ensure effective communication and better patient experiences.

One theme highlighted by survey respondents was the use of interpreters or translators to address language discordance. Interpreters can provide an effective way of improving the communication between a healthcare providers and patients. Indeed, the use of professional interpreters can decrease the risk of adverse events (37). The use of unofficial interpreters such as bilingual healthcare staff, or family members, are also essential in reducing linguistic barriers (38,39). However, reliance on untrained interpreters may lead to issues, particularly concerning the precision of medical vocabulary (40). Ultimately, using trained interpreters improves precision and decreases errors. This emphasizes the importance of understanding the language that is used in a medical context so that healthcare providers might better understand a problem presented by a patient.

Approximately one-third of respondents from higher percentage Welsh-speaking areas (32%) report feeling restricted by their inability to communicate in Welsh or bilingually. This is consistent with a survey of pharmacy users in Wales, where 54% of respondents agreed that they struggled to think of English word(s) when explaining their concerns to a pharmacist (36). Notably, Roberts et al (2003) report that Welsh and English speakers express their pain differently. Welsh speakers often use metaphors and pairs of descriptors to convey their experiences more effectively (22). Furthermore, 38% of the terms used by Welsh speakers were absent from the standard English McGill Pain Questionnaire (41), a tool used in clinical practice to generate a quantitative figure for ‘pain’ using a patients verbal descriptors (22). This evidence highlights the limitations of English-only healthcare for bilingual Welsh patients and underscores the importance of offering bilingual services to support effective communication and patient care.

There are studies suggesting that care in a patients first language may also have a beneficial effect on a patient’s physical health. Fernandez et al (2012) showed that limited English proficiency contributes to poorer glycemic (HbA1C) management among diabetic Latino patients in the U.S.A. (7). Furthermore, glycemic management improved when Latino patients switched from an English only to a Spanish-speaking physician (42). Language concordance also improves adherence of Latino asthma patients to their treatment regimes, which were in the context of asthma (16) and cardiovascular disease (43). In the Welsh context, Owen & Morris (2012) reported that Welsh-speaking patients treated by non-Welsh speaking physiotherapists had lower functional outcome measures compared to those treated by bilingual therapists (44). Thus, language concordance can impact both physical and mental wellbeing (24), emphasizing a need for further evidence on the physical impact in the Welsh context.

The majority of our survey respondents expressed dissatisfaction with having to forgo their right to use their preferred language for faster healthcare access. However, they acknowledged challenges faced by the NHS, including staffing shortages, recruitment difficulties, and a lack of bilingual healthcare professionals. Recruiting and retaining GPs, particularly in rural areas is challenging (45). Verma et al. (2016) suggests strategies like financial incentives to recruit and retain GPs to the areas where demand is greatest (46). Despite the WLS set by the Welsh language commissioner, Prys and Matthews (2023) reported that the Welsh language is largely absent from local health boards’ well-being objectives, limiting the realistic implementation of bilingual care in Wales (47). However, the Welsh Government strategy *’More than just words’*, re-released in 2022, describes bilingualism as being important for patient-centred care and outlines initiatives to increase the Welsh ‘active offer’ in healthcare (48).

To facilitate the ‘active offer’ of Welsh language provision, patients could indicate their language preference on medical records, which was supported by most respondents. However, accessibility challenges remain, as suggested by one respondent, *“Considering the fact that other doctors (some of them non-Welsh speaking) need to read notes, we would like to see a system where any doctor would be able to read the notes in either language”* (Table 3). Alternatively, a formal UK protocol should be devised on communicating safely and effectively when patients or clinicians use a second language, as has been investigated with Chinese patients in Australia (49). However, for some it is not a matter of choice but of necessity, as they do not have a confident grasp of English (24). For example, bilingual people with dementia may lose fluency in their second language (50). In such cases, assessments should be conducted in both languages, with the best result accepted as most accurate (51). Although many Welsh speakers are fluent in English, research indicates that they may struggle to fully express themselves in their second language (22,24,52,53). This raises the question of how (and how well) the first-language Welsh speakers process, understand and interpret English-medium information and guidance.

Clearer guidance is needed to provide effective healthcare for speakers of minority languages. In the Basque Country, bilingual pathways have been established, starting in primary care, where GPs are responsible for raising awareness about patients’ language preferences before they access specialist care (54). Investigating and adapting successful approaches from other minority language contexts will be vital for developing a robust framework for bilingual healthcare in Wales. In this study we have shown that Welsh speakers value bilingual healthcare, however, many have not been offered the option to communicate in Welsh and there is a feeling that their language needs are not prioritized in primary care. The current disparity in bilingual service availability potentially impacts patient comfort, communication, and quality of care, thus there is a need for increased awareness, active offers, and improved rights to support communication in a patients’ preferred language.

## Data Availability

All relevant data are within the manuscript and its Supporting Information files.

## Notes

### Competing Interest Statement

The authors have declared no competing interest.

### Funding Statement

The author(s) received no specific funding for this work.

### Author Declarations

Ethical approval was granted by Swansea University Medical School Research Ethics committee, (reference number 2021-0089, 18/11/2021).

